# Implementing intravenous iron for maternal anemia in Nigeria: A qualitative study of healthcare provider experiences using the Normalization Process Theory

**DOI:** 10.1101/2025.11.06.25339710

**Authors:** Damilola Onietan, Chisom Obi-Jeff, Yusuf Adelabu, Mobolanle Balogun, Opeyemi Akinajo, Esther O. Oluwole, Aduragbemi Banke-Thomas, Bosede B. Afolabi, Ejemai Eboreime

## Abstract

**Background:** Maternal anemia remains a significant public health challenge in Nigeria, affecting approximately 25-46% of pregnant women and contributing to adverse maternal and neonatal outcomes. Intravenous (IV) iron provides a promising alternative to conventional oral iron supplementation for managing moderate to severe maternal anemia; however, its implementation in resource-limited settings faces numerous challenges. This study aimed to understand how the implementation of IV iron became embedded in everyday practice, including factors influencing skilled healthcare providers’ (SHPs) engagement, workflow integration processes, and sustainability.

**Methods:** This was a qualitative study embedded within the Implementation Research for Intravenous Iron Use in Pregnant and Postpartum Women in Nigeria (IVON-IS) project across six healthcare facilities in Lagos, Nigeria. Eighteen key informant interviews were conducted with purposively sampled SHPs across the six IVON-IS facilities. The data were analyzed deductively based on the Normalization Process Theory (NPT) constructs (coherence, cognitive participation, collective action, and reflexive monitoring), and inductively to identify themes related to each of these constructs.

**Results:** Our study revealed strong coherence among SHPs regarding the purpose and benefits of IV iron compared to traditional treatments. Cognitive participation varied across facilities, with leadership support and patient-centred motivation emerging as critical facilitators. Collective action was faced with challenges, including workflow disruptions, staffing constraints, and space limitations, despite adequate resource provision. Reflexive monitoring processes were robust, with providers continuously evaluating effectiveness through clinical outcomes and patient feedback while expressing concerns about long-term sustainability.

**Conclusion:** The implementation of IV iron for maternal anemia in Nigeria demonstrated variability across NPT constructs, with strong coherence and reflexive monitoring but challenges in cognitive participation and collective action. Critical success factors included strong leadership support, adequate resource provision, continuous quality improvement processes, and proactive sustainability planning. These findings provide valuable guidance for scaling up IV iron use as part of comprehensive maternal health services in Nigeria and similar resource-constrained settings.

## Introduction

Maternal anemia remains a significant public health challenge worldwide, contributing significantly to maternal and perinatal mortality and morbidity(1). Globally, it affected 500 million women of reproductive age 15–49 years in 2019; out of which, 32 million were pregnant women. The burden is particularly severe in Sub-Saharan Africa (SSA), with over 40% of pregnant women affected with anemia(2,3). Limited healthcare resources, nutritional deficiencies, and infectious diseases create a complex web of risk factors that perpetuate high prevalence rates in SSA(4).

Nigeria, the most populous country in Africa, exemplifies this crisis, with maternal anemia affecting an estimated six in ten pregnant women(5),(6). Iron deficiency accounts for 25-46% of these anemia cases, resulting from inadequate nutritional iron intake, increased iron requirements for placental and fetal development, maternal erythropoiesis, and preparation for blood loss during delivery(6,7). Other causes of anaemia in Nigeria include nutritional disorders, malaria, infections, hemoglobinopathies, and chronic diseases(8). These various types of anaemia during pregnancy can result in adverse maternal and neonatal outcomes. In mothers, consequences may include postpartum haemorrhage, fatigue, and an elevated risk of postpartum depression. Newborns are also at higher risk for low birth weight, preterm birth, stillbirth, and neonatal death(7,9).

Anemia in pregnancy is an independent risk factor for postpartum anaemia (PPA) as iron stores tend to remain low for several months after childbirth. This prevalence is highest in low and middle-income countries with prevalence estimated to be about 50-80% in postpartum women (10,11). Despite its high prevalence, postpartum anemia may not be diagnosed on time. Other risk factors of PPA include young maternal age, pregnancy anaemia, postpartum haemorrhage, inadequate antenatal care visits, type of birth, and poor adherence to iron and folic acid supplementation in pregnancy(12). The consequences of PPA include tiredness, breathlessness, palpitations, reduced cognitive abilities, emotional instability, distress, increased risk of infections, and postpartum depression(13).

Traditional approaches to managing maternal anemia in Nigeria have mainly depended on oral iron supplementation (120 mg of elemental iron daily until hemoglobin rises to normal). However, oral iron typically takes several months to correct anemia and replenish iron stores. Additionally, 50% of patients experience gastrointestinal adverse effects, including vomiting, constipation, diarrhea, and abdominal pain, all of which contribute to poor adherence(14). In Nigeria, compliance rates with oral iron supplementation are estimated to be as low as 65%(15). “Blood transfusion is used in severe cases but carries significant risks, including transfusion reactions and transmission of infections. Additionally, blood supply shortages constrain its availability and use(5).

Intravenous (IV) iron represents an alternative approach for treating moderate to severe maternal anemia. Modern IV iron formulations such as ferric carboxymaltose (FCM), allow for rapid administration of high doses in a single infusion with minimal risk of adverse reactions and more rapid improvement in hemoglobin levels compared to oral iron(16). A single dose of FCM can be infused in 15 minutes, rapidly replenishing iron stores, and has been demonstrated to be cost-effective compared to IV sucrose(17). A randomised-control clinical trial in Nigeria demonstrated that FCM is safe, with no serious drug-related adverse events reported among 527 pregnant women(16).

While IV iron is widely used in high-income settings, its implementation into routine care in resource-limited settings, such as Nigeria and Malawi, has been limited by factors including cost, availability, healthcare system constraints, and misconceptions about treatment modalities(18,19). A recent qualitative study examining the acceptability of IV iron among pregnant women, domestic decision-makers, and healthcare providers in Nigeria revealed positive attitudes but identified barriers, including infrastructure constraints, training needs, and economic challenges(18). While the research on acceptability provides important foundational knowledge, significant gaps remain in understanding how IV iron becomes embedded into routine clinical practice when transitioning from controlled trial settings to real-world implementation. Specifically, previous studies have not adequately addressed how healthcare providers institutionalised IV iron into their routine practice, factors influencing provider engagement, workflow integration processes, and sustainability planning approaches.

Understanding how complex healthcare interventions become embedded in everyday practice is crucial for translating evidence into sustainable improvements in maternal health outcomes. To address this gap, our research objectives are to understand how healthcare providers make sense of IV iron as a treatment option for maternal anemia; identify factors influencing provider engagement with IV iron implementation; examine how IV iron administration was operationalized within existing clinical workflows; and analyze the processes through which providers evaluated and refined the implementation.

## Methods

### Study design

This is an exploratory, qualitative implementation study using framework analysis. The study was conducted within the Implementation Research for Intravenous Iron Use in Pregnant and Postpartum Women in Nigeria (IVON-IS) project(20,21). The IVON-IS project was a multi-site implementation study conducted in Lagos, Nigeria, aimed at evaluating the feasibility, acceptability, and effectiveness of introducing IV iron (FCM) as a routine treatment option for pregnant and postpartum women with moderate to severe anemia(21). We report this study following the consolidated criteria for reporting qualitative research (COREQ)(22). (See additional file 1)

### Study setting

The study was conducted in Nigeria, a country characterized by significant diversity, encompassing 36 states, a Federal Capital Territory, 774 local government areas, over 400 ethnic groups, and approximately 450 languages across its six geopolitical zones(23). The Nigerian health system is delivered across three tiers: primary, secondary, and tertiary. This study was conducted in a cluster of six healthcare facilities as part of the IVON-IS project in Lagos State, South-West Nigeria(21). The facilities included one tertiary hospital (Federal Medical Center Ebute Metta – designated as TF); two secondary hospitals (Harvey Road General Hospital – SHF1 and Ebute Metta General Hospital – SHF2); one comprehensive primary healthcare center (Akerele Health Center – designated as PHC); and two private hospitals, Havana Specialist Hospital – PHF1 and Kensington Adebukunola Adebutu Foundation (KAAF) Maternity Hospital – PHF2. These facilities were purposively selected to represent different levels of the Nigerian healthcare system while maintaining geographic proximity within the state’s established referral network. The selected healthcare facilities collectively serve a diverse patient population, providing prenatal and postnatal care services to women from various socioeconomic and cultural backgrounds.

### Recruitment procedure

We aimed for a sample size that considered information power based on the study aim, sample specificity, involvement of different cadres of SHPs across various health facilities, and the high quality of dialogue from multiple perspectives(24). We employed a purposive sampling technique to identify skilled healthcare providers involved in the IVON-IS project, who were then recruited for this study. We invited 18 SHPs, three from each facility, and none declined. Of the 18 recruited SHPs, 13 were physicians and five were nurses, across the six implementing health facilities. SHPs are defined as competent maternal and newborn health (MNH) personnel who can provide effective, uninterrupted, high-quality care(25).

### Data collection

Three trained interviewers facilitated key informant interviews (KIIs) with SHPs involved in the IVON-IS project. KIIs were selected as they provided opportunity for in-depth discussions and are practical for healthcare providers(26). Each interview involved two team members, one to conduct the interview and another to monitor and record.

The KIIs were conducted with semi-structured topic guides. The guides were developed using NPT constructs while allowing for emergent themes. Interview questions explored providers’ understanding of IV iron’s purpose and benefits (coherence); their willingness to adopt IV iron and factors motivating participation (cognitive participation); workflow integration and resource allocation processes (collective action); and evaluation methods and sustainability planning (reflexive monitoring).

Interviews were conducted in English at the six health facilities between October 23 and December 5, 2024 (Table 1). Each interview lasted between 30-45 minutes. Topic guides were piloted prior to data collection, using similar participants and adjustments made for clarity. All interviews commenced after obtaining consent from participants. All interviews were audio-recorded with participant consent and transcribed verbatim.

**Table 1.**
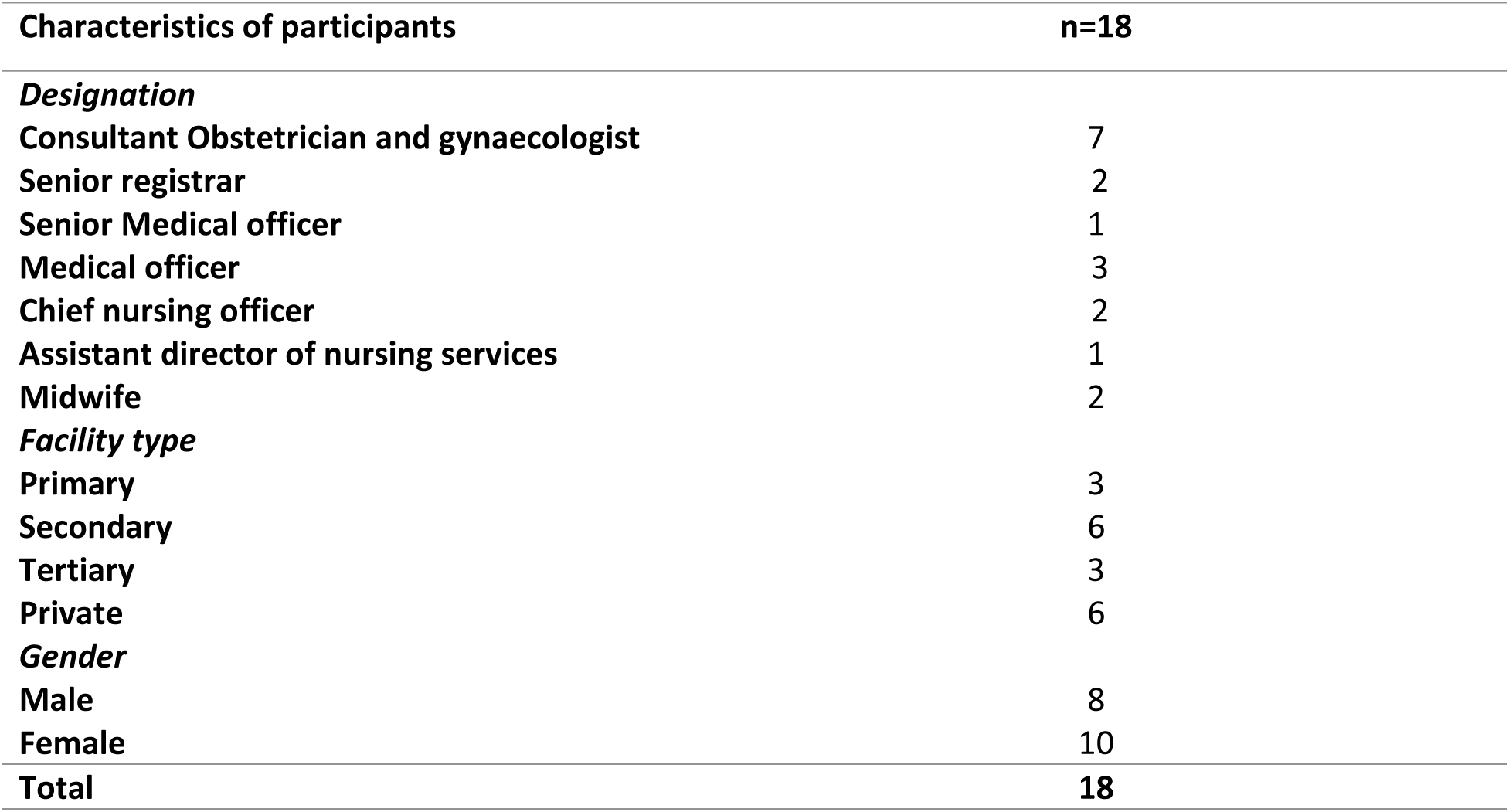
Summary profiles of participants interviewed.

| Characteristics of participants | n=18 |
| --- | --- |
| ***Designation*** |  |
| Consultant Obstetrician and gynaecologist | 7 |
| Senior registrar | 2 |
| Senior Medical officer | 1 |
| Medical officer | 3 |
| Chief nursing officer | 2 |
| Assistant director of nursing services | 1 |
| Midwife | 2 |
| ***Facility type*** |  |
| Primary | 3 |
| Secondary | 6 |
| Tertiary | 3 |
| Private | 6 |
| ***Gender*** |  |
| Male | 8 |
| Female | 10 |
| **Total** | **18** |

### Theoretical framework

This study is grounded in the Normalization Process Theory (NPT), which views healthcare practices as fundamentally social phenomena constructed through collective work rather than implemented through top-down directives(27,28). The NPT’s ontological position acknowledges that IV iron implementation entails complex negotiations among healthcare providers, organizational contexts, and existing care practices, thereby positioning implementation as an emergent social process rather than a predetermined technical pathway. The NPT adopts a constructivist approach that prioritizes healthcare providers as active agents who interpret, adapt, and sustain new practices within their respective contexts. This aligns with our research objectives to understand how providers make sense of IV iron, engage with its implementation, and operationalize it within existing workflows. The NPT examines normalization through four core constructs: **Coherence** (how individuals understand the intervention and differentiate it from existing practices); **Cognitive participation** (how individuals commit to and engage with the intervention); **Collective action** (how the intervention is operationalized through coordinated work); and **Reflexive monitoring** (how individuals evaluate and refine the intervention). The NPT’s focus on these normalization processes is particularly relevant for resource-constrained settings, where successful integration requires fundamental shifts in how maternal anemia care is conceptualized and delivered through coordinated efforts across multiple levels of the healthcare system(27,28).

## Data analysis

The NPT framework was used to inform and guide the analysis. During data analysis, the four NPT constructs (coherence, cognitive participation, collective action, and reflexive monitoring) served as initial coding categories, with subcategories developed inductively from the data. This approach facilitated a systematic examination of implementation processes while remaining open to context-specific factors that might not be explicitly captured by the theory. The analytical process involved mapping participant responses to NPT constructs, identifying patterns across facilities, and exploring relationships between theoretical components to understand how IV iron became normalized (or faced barriers to normalization) within the healthcare settings. Next, two qualitatively trained researchers (DO and EE) randomly selected two transcripts and coded them independently. The codes were discussed, and a final coding framework was developed. EE then coded the remaining data. All codes were subsequently organized into categories, after which key themes and sub-themes emerged from the data(29). Analysis was conducted iteratively, with researchers identifying themes related to each NPT construct and examining the relationships between these themes to understand the overall normalization process. NVivo 12 Plus (QSR International, Memphis, USA) aided analysis.

## Ethical declarations

### Ethics approval and consent to participate

Ethical approval was obtained from the Health Research and Ethics Committees of the Lagos University Teaching Hospital (ADM/DCST/HREC/APP/5328) and Federal Medical Centre, Ebute Metta (HREC 22-22). Operational approval was obtained from the Lagos State Health Service Commission and the Lagos State Primary Health Care Board. In addition, all participants were informed about voluntary participation, privacy, confidentiality, risks, and benefits. Verbal consent and permission for audio recording were obtained from all participants before conducting interviews.

## Results

Our findings revealed that the implementation of IV iron followed a progressive pathway, with SHPs moving through sequential yet interconnected stages from understanding the intervention to ensuring its sustainability. The following sections present detailed findings organized by the four NPT constructs.

### Coherence: Making sense of IV Iron

The SHPs demonstrated a strong understanding of IV iron’s purpose, benefits, and differences compared to traditional treatments for anemia. This understanding evolved from initial uncertainty to clear conceptualisation throughout the implementation process. Providers conceptualised IV iron as both a treatment option and an innovation in maternal healthcare delivery, frequently describing it as a "novel method of treatment" for reducing the burden of moderate to severe iron deficiency anemia. As a male SHP articulated,

> “It serves to reduce the burden for iron deficiency anaemia among pregnant and postpartum women who have moderate to severe anaemia." **Senior Medical Officer, Male, SHF2**

The SHPs differentiated it from existing practices, primarily emphasizing its key advantage of single-dose administration versus daily oral medication.

> "*The previous method [oral iron] involved women taking medication* every *day; meanwhile, administration of IV iron involves administration of a single dose of iron*."

This understanding was consistently echoed by SHPs across different facilities.

> "*The purpose of using IV-ion for maternal anaemia is to help correct moderate to severe anaemia in order to improve maternal well-being and also foetal well-being too… So that the mother can have an optimal blood level, in other words, an optimal haemoglobin level during pregnancy and even immediately after delivery*." **Medical Officer, Male, PHC**

Multiple benefits were identified by the SHPs that reinforced IV iron’s value in clinical practice, including improved patient compliance, efficacy for patients with asymptomatic severe anemia, and reduced treatment burden. Providers also recognized broader socioeconomic benefits, particularly the cost savings for patients who might otherwise require blood transfusion at secondary facilities.

> "The main purpose is to reduce anemia in pregnancy and postpartum women, and it has been very, very helpful because if they are being referred to the secondary facility they need to pay for blood transfusion, they need to pay for some other logistics but here the purpose was fulfilled it is free, its safe and it is accessible**." Chief Nursing Officer, Female, PHC**

This comprehensive understanding of purpose, distinctiveness, and benefits created a strong foundation for implementation.

### Cognitive participation: Engaging with IV iron implementation

The analysis revealed varying engagement levels among SHPs with important insights into motivational factors and the role of key individuals in driving adoption. Leadership support emerged as a critical facilitator, with facility management creating an enabling environment for the intervention.

> "We benefitted from the buy-in from management… the fact that the medical director, we had the full support of the medical director, it really helped**." Senior Medical Officer, Male, SHF2**

This support was operationalized through Implementation Management Teams (IMTs) that coordinated activities, conducted regular monitoring, performed data analysis, and produced reports. Within each facility, specific local champions drove implementation forward by maintaining momentum and addressing challenges. A nurse midwife described this leadership involvement:

> "Our apex nurse is part of the IMT… And our Medical director, too, was aware of it… So, they all supported." **Chief Nursing Officer, Female, PHC**

Staff engagement varied considerably across facilities and over time. Initial resistance gradually evolved into acceptance as SHPs became more familiar with the intervention. The implementation typically followed a pattern of gradual adoption, characterized as "slow initially" but eventually becoming "seamless" after persistent counselling and education efforts. Several factors motivated SHPs’ engagement, including professional interests, personal values alignment, and financial incentives, which proved especially important for lower-cadre staff. Despite these varied motivations, patient benefit consistently emerged as the primary driver of engagement. As a female nurse emphasized,

> "My patient is going to benefit much from it… so that’s one of the main reasons." **Assistant Director of Nursing Services, Female, SHF2**

> "The factors that motivated us to say ok, let us find a lasting solution to this If IV-ion is the solution, then let’s go ahead and use it to treat maternal anaemia**." Medical Officer, Male, PHC**

According to the SHPs, the patient-centred motivation helped sustain commitment when implementation challenges arose.

### Collective action: Operationalizing IV iron use

SHPs described how IV iron was integrated into existing practices, including workflow adaptations, resource utilization, and addressing implementation challenges. Clinical workflow integration required significant adaptations, including securing an appropriate physical space for administration and modifying screening protocols to test all antenatal patients for anemia on a routine basis.

> "*Now we* routinize *the screening so all ante-natal patients are now tested for anaemia because we are trying to be on the lookout for those that have moderate or severe anaemia*." **Medical Officer, Male, SHF1**

Furthermore, the SHPs emphasized that integrating IV iron into existing workflows necessitated a coordinated effort across multiple units and leveraging existing meeting structures.

> "*We have the* pharmacy*, we have the Lab, we have the Nurses, we have the Doctors. We all work hand in hand. If there are any challenges, we have a meeting every month. We discuss our challenges, what ways we can use to improve, and assist with the project… Everybody has their own role to perform*." **Consultant Obstetrician and Gynaecologist, Female, SHF1**

Staff roles were redistributed, with specific responsibilities assigned for screening, consent processes, and administration. Physicians typically assumed primary responsibility for IV iron administration, creating new collaborative arrangements that required coordination among diverse SHPs.

Resource availability significantly facilitated implementation, with providers highlighting the importance of having appropriate equipment and supplies. This resource provision minimized logistical barriers to implementation. One male provider noted,

> "*Everything was in place from the HemoCue to diagnosis; quickly, we were able to map out processes to make it work in this facility.*" **Medical Officer, Male, SHF1**

Despite careful planning, several implementation challenges emerged. Staffing constraints affected service delivery, with the additional workload impacting existing clinical duties. A female provider described this impact:

> "*It affected my daily work routine because it makes me leave the patient I am attending to, whether in the gynaecology clinic or the antenatal clinic*." **Assistant Director of Nursing Services, Female, PHF2**

This was echoed by a nurse midwife who noted:

> "*It has affected my work routine mostly because of the ‘japa’ [brain drain]; most of the nurses have travelled out [of the country], so the workload is much. At times, the doctors might be busy downstairs. We need to pick their line and set their lines for them*." **Chief Nursing Officer, Female, PHC**

Space limitations and logistical issues also presented obstacles that required creative problem-solving.

Patient acceptance challenges were also noted, particularly related to the intervention’s novelty, with literacy levels and communication barriers complicating patient engagement. A medical officer shared:

> "*Only one patient complained that she didn’t want IV iron. We counseled her, and she said No. And then I think we tried to speak with the husband, but we couldn’t get through to him. That was the only patient I know of that declined the infusion*." **Medical Officer, Female, PHC**

### Reflexive monitoring: Evaluating IV iron implementation

SHPs continuously assessed IV iron’s effectiveness, addressed emerging issues, and gathered patient feedback to refine the implementation process while considering sustainability challenges.

Some SHPs reported that they primarily measured effectiveness by tracking clinical outcomes, especially improvements in hemoglobin levels. Some providers noted discrepancies between expected and observed clinical responses, with hemoglobin levels sometimes taking longer to rise than anticipated. As one male stated:

> "We were expecting that within two to three weeks we should start seeing the rise, but most times we don’t even see any rise at all till the third and fourth week." **Medical Officer, Male, SHF1**

These observations prompted adjustments to patient counselling regarding expected outcomes.

SHPs stated that regular evaluation meetings facilitated continuous quality improvement through systematic bottleneck analysis and data-driven decision-making.

> "We usually have meetings to analyse for bottlenecks… so that we could identify the challenges and how to overcome them." **Doctor, Female, TF**

Implementation teams used these processes to identify and address emergent issues, including knowledge gaps among new staff and clinical errors, which became learning opportunities.

Patient feedback was generally positive, regarding the intervention. A nurse midwife noted:

> "*They are always very happy o! They come back saying* thanking *us, ’nurse thank you o’; and the good thing was that it was free. They are so happy because they don’t have to pay for it*." **Nurse Midwife, Female, PHF1**

This appreciation extended to word-of-mouth promotion among patients and positive reception of comprehensive anemia screening. One SHP described:

> "*They are happy, they want to tell* their *other friends, ‘Go to that hospital, they will give you’. You know, because we attend to a lot of semi-literate, let me say that, so when they come and they see good outcomes, they want to bring their friends*. **"Medical Officer, Male, PHC**

Despite the successful implementation, participants expressed significant concerns about the long-term sustainability, particularly regarding access to the IV iron after the project end. A medical director voiced this anxiety:

> *"My greatest fear now is that now that* project *is rounding up… I don’t know if we will still have access to some of these drugs." **Medical Director, Female, PHF1***

A nurse midwife similarly expressed concern:

> "*When the project was on, it has been* our *routine care, but now that the project has stopped and no [more] support, how do we do it? I cannot use my salary to buy the injection. I cannot use my salary to buy all the consumables**." Assistant Director of Nursing Services, Female, PHF2***

SHPs demonstrated proactive engagement with sustainability planning through institutional advocacy efforts and proposing potential solutions focused on resource availability and cost reduction strategies. A medical officer described their efforts:

> "*Now we are trying to have a meeting with the Medical officer of Health, that is the head of the clinical service in the local government, and then the other stakeholders like the special adviser on health and some other stakeholders in the health department of the local government, in addition to primary health board to see if there can be some kind of incentives and support." **Medical officer, Male, PHC***

Suggestions for sustaining the intervention included seeking Non-Governmental Organisation (NGO) sponsorship, implementing a drug-revolving system with government subsidies, and incorporating patient co-payments. These sustainability planning efforts underscore the providers’ commitment to maintaining IV iron as part of routine care, despite the challenges of transitioning from a donor-supported project to a self-sustaining program.

## Discussion

This study applied NPT to analyze the implementation of IV iron for the treatment of maternal anemia in the selected healthcare facilities in Lagos state, revealing important insights into the normalization process. The study aimed to understand how IV iron was instutionalized into everyday practice by SHPs. Our findings demonstrate that the successful implementation of IV iron required not only technical knowledge and resources but also careful attention to the social and organizational contexts in which the intervention was introduced.

The strong coherence observed among SHPs suggests that efforts, such as training of the SHPs on the safe administration of IV iron, to build a shared understanding of IV iron’s purpose and benefits, were largely effective. This finding aligns with that of the previous research, highlighting the importance of collective sense-making by SHPs as a foundational element for the effective implementation of new interventions(18). The clear differentiation between IV iron and existing treatments, particularly emphasizing single-dose administration versus daily oral medication, helped to establish its value proposition for both providers and patients. According to the providers across facilities, this understanding encompassed both maternal and fetal benefits, creating a compelling rationale that supported adoption.

Leadership support was identified as the primary facilitator of strong cognitive participation. This finding resonates with implementation science literature, which emphasizes the role of organizational leadership in creating environments conducive to change(30). The gradual pattern of adoption observed from initial resistance to eventual acceptance underscores the importance of sustained engagement efforts throughout the implementation process. Financial incentives proved important, particularly for lower-cadre staff, though the predominance of patient benefit as a motivating factor suggests that framing interventions in terms of their impact on patient outcomes may enhance healthcare worker engagement beyond material incentives. As one provider noted, ‘’they redistributed financial incentives to ensure broader staff motivation”, highlighting the importance of addressing perceived inequities in incentive distribution.

Collective action faced significant challenges despite adequate resource provision. The workflow disruptions and staffing constraints experienced highlight the importance of considering how new interventions interact with existing systems and practices.(31) The SHPs demonstrated considerable adaptability in addressing these challenges, such as relocating laboratory personnel to antenatal clinics to streamline screening processes and redistributing tasks among available staff. These adaptive responses reflect the dynamic nature of implementation and the agency of local actors in shaping how interventions are operationalized. The implementation was further facilitated by the availability of appropriate equipment, particularly the HemoCue device, which enabled immediate point-of-care hemoglobin test. The challenges with staffing were exacerbated by the broader healthcare workforce issues in Nigeria, referred to by one provider as "jápa" (a colloquial term for emigration of healthcare workers), highlighting how macro-level healthcare system constraints can impact facility-level implementation. These findings suggest that implementation planning should include detailed workforce analysis to identify potential bottlenecks and resource requirements before introducing new interventions.

However, reflexive monitoring processes were robust, with continuous evaluation and improvement mechanisms in place. The discrepancies noted between expected and observed clinical responses highlight the importance of setting realistic expectations and providing clear information to both providers and patients about intervention outcomes. Regular ‘bottleneck analysis’ meetings facilitated shared learning and problem-solving, enabling adaptation of implementation strategies as challenges emerged. Patient feedback served as a crucial mechanism for evaluating implementation success, with positive outcomes and experiences driving continued engagement from both patients and SHPs.

The concerns expressed about long-term sustainability emphasize the need for early attention to sustainability planning, particularly in resource-constrained settings where external funding may be time-limited. SHPs in our study demonstrated proactive engagement with sustainability planning, proposing innovative financing mechanisms such as drug revolving funds and public-private partnerships. However, these proposed solutions face significant challenges in a context where patients’ ability to pay is limited and government resources are constrained. This tension reflects broader challenges in scaling and sustaining evidence-based interventions in low-resource settings.(32)

Our findings offer valuable insights into the implementation of IV iron therapy for maternal anemia in resource-constrained settings and contribute to understanding how healthcare innovations become integrated into practice, particularly in these settings. They highlight the interplay between intervention characteristics, implementation strategies, provider motivation, and organizational context in shaping implementation outcomes. The successful integration of IV iron into maternal care routines during the project period demonstrates the potential for embedding this intervention into standard care. However, sustainability concerns emphasize the need to address financial and systemic barriers to long-term adoption. constructs represent core, universal facilitators for implementation success in settings constrained by limited resources. As in other applications of NPT in low-resource settings, we note similarities in the importance of leadership support, resource availability, and alignment with existing staff values(33). However, our study uniquely highlights the role of financial considerations, both incentives for providers and costs for patients, in shaping implementation processes and sustainability. Unlike implementations in high-resource settings, providers in our study faced substantial challenges related to basic infrastructure, staffing, and supply chain management, requiring greater adaptability and innovation. Future research should explore patient perspectives on IV iron, examine the long-term sustainability of implementation, and investigate the effectiveness of different financing mechanisms for ensuring continued access to IV iron beyond the initial project.

### Strengths and Limitations

The use of the NPT offered the possibility to understand how a new intervention, IV iron, was instutionalized in the health facilities from the perspective of SHPs. In addition, the sample size recruited for the study, the appropriateness of which was assessed with the principle of information power, offered us an opportunity to have comprehensive narratives that enabled data and analytical sufficiency. Additionally, to enhance trustworthiness, a transparent process was maintained throughout the study, and a rigorous analytical approach was employed to ensure credibility and promote reflexivity. Despite these strengths, our study has some limitations. First, it relied on KIIs with SHPs, potentially missing perspectives from patients and health system administrators. Secondly, our data collection occurred during the active implementation phase, and we were unable to assess the long-term sustainability of the intervention beyond project completion.

### Recommendations for Implementation

Based on our findings, we propose the following recommendations to facilitate the successful implementation of IV iron in similar resource-constrained settings:

1. **Develop comprehensive educational materials**: Create educational resources for both SHPs and patients that clearly explain IV iron’s mechanism of action, benefits, and expected outcomes. These materials should address common misconceptions and set realistic expectations regarding the timeline for hemoglobin improvement.
2. **Secure early leadership engagement**: Formally engage facility leadership from the planning phase to establish champions who can facilitate institutional buy-in. Leadership should be involved in creating dedicated implementation teams with clear roles and responsibilities.
3. **Perform pre-implementation workflow analysis**: Before introducing IV iron, conduct a detailed analysis of existing clinical workflows to identify potential integration points and resource needs. This should include space allocation, staffing requirements, and scheduling considerations to minimize disruption to existing services.
4. **Establish structured monitoring systems**: Implement regular data collection and review procedures to track clinical outcomes, patient experiences, and implementation challenges. This should include standardized documentation tools and scheduled review meetings to address emerging issues promptly.
5. **Develop sustainability planning from inception**: Create sustainability plans at the outset that address long-term financing, resource procurement, and integration into existing health system structures. Engage with policymakers early to advocate for inclusion of IV iron in national guidelines and essential medicine lists.
6. **Design tailored training programs**: Develop role-specific training that addresses not only technical aspects of IV iron administration but also strategies for patient counseling, consent processes, and management of potential adverse effects. Training should be ongoing to accommodate staff turnover.
7. **Establish clear communication channels**: Create formal communication structures between different cadres of SHPs involved in maternal anemia management to ensure coordination and information sharing about patient care.
8. **Frame intervention around patient benefits**: Emphasize concrete patient benefits when communicating about IV iron to healthcare workers to leverage patient-centered motivation as a driver of engagement.
9. **Explore innovative financing mechanisms**: Investigate approaches such as drug revolving funds and public-private partnerships to ensure financial sustainability beyond the initial project funding.

## Conclusion

This study demonstrates that successfully implementing IV iron treatment for maternal anemia in resource-limited settings, such as Nigeria, requires more than just training healthcare workers; it demands comprehensive changes to the health system, including updated national policies, sustainable financing mechanisms, and restructured service delivery models that can integrate new treatments without disrupting existing care. When these broader contextual factors are addressed alongside technical implementation, IV iron therapy has significant potential to reduce maternal anemia and improve outcomes for mothers and babies in challenging healthcare environments.

## List of abbreviations

IV: Intravenous iron
SHP: Skilled healthcare providers
IVON-IS: Implementation Research for Intravenous Iron Use in Nigeria
NPT: Normalization Process Theory
SSA: Sub-Saharan Africa
PPA: Postpartum anemia
FCM: Ferric Carboxymaltose Maltose
COREQ: Consolidated criteria for reporting qualitative research
TF: Tertiary facility
SHF1: Secondary health facility 1
SHF2: Secondary health facility 2
PHC: Primary healthcare facility
PHF1: Private healthcare facility 1
PHF2: Private healthcare facility 2
MNH: Maternal and Newborn Health
LMICs: Low- and Middle-Income Countries
KII: Key informant interview
IMT: Implementation Management Team
NGO: Non-governmental organisation
HREC: Health Research and Ethics Committee

## Supporting information

S1 File. COREQ Checklist

## Data Availability

All relevant data are within the manuscript and its Supporting Information files. Audio files are ethically restricted.

## Acknowledgment

The authors sincerely thank all the SHPs and the IMTs for their valuable contribution to this project.

## Author’s Contribution

EE conceived the research idea. DO led the project management and implementation. DO led data collection. DO and EE coded the data set. EE and DO performed and interpreted the data analysis. BBA obtained funding. DO prepared the initial draft of the manuscript. DO, EE, BBA, ABT, COJ, YA, MB, EOO, and OA critically revised the manuscript. All authors endorsed and contributed to the last version of the manuscript for publication

